# Modeling Pulmonary Cystic Fibrosis in a Human Lung Airway-on-a-chip

**DOI:** 10.1101/2021.07.15.21260407

**Authors:** Roberto Plebani, Ratnakar Potla, Mercy Soong, Haiqing Bai, Zohreh Izadifar, Amanda Jiang, Renee N. Travis, Chaitra Belgur, Mark J. Cartwright, Rachelle Prantil-Baun, Pawan Jolly, Sarah E. Giplin, Mario Romano, Donald E. Ingber

**Affiliations:** Wyss Institute for Biologically Inspired Engineering at Harvard University, Boston, MA; Center on Advanced Studies and Technology (CAST), Department of Medical, Oral and Biotechnological Sciences, “G. d’Annunzio” University of Chieti-Pescara, Chieti, Italy; Vascular Biology Program and Department of Surgery, Boston Children’s Hospital and Harvard Medical School, Boston, MA; Harvard John A. Paulson School of Engineering and Applied Sciences, Cambridge, MA

**Keywords:** Cystic Fibrosis, Neutrophils, Pseudomonas, Organ Chip, Microfluidics

## Abstract

**Background:** Cystic fibrosis (CF) is a genetic disease caused by mutations in the gene encoding the cystic fibrosis transmembrane conductance regulator (CFTR), which results in impaired airway mucociliary clearance, inflammation, infection, and respiratory insufficiency. The development of new therapeutics for CF are limited by the lack of reliable preclinical models that recapitulate the structural, immunological, and bioelectrical features of human CF lungs.

**Methods:** We leveraged organ-on-a-chip technology to develop a microfluidic device lined by primary human CF bronchial epithelial cells grown under an air-liquid interface and interfaced with pulmonary microvascular endothelial cells (CF Airway Chip) exposed to fluid flow. The responses of CF and healthy Airway Chips were analyzed in the presence or absence of polymorphonuclear leukocytes (PMNs) and the bacterial pathogen, *Pseudomonas aeruginosa*.

**Results:** The CF Airway Chip faithfully recapitulated many features of the human CF airways, including enhanced mucus production, increased cilia density and a higher ciliary beating frequency compared to chips lined by healthy bronchial epithelial cells. The CF chips also secreted higher levels of IL-8, which was accompanied by enhanced PMN adhesion to the endothelium and transmigration into the airway compartment. In addition, CF Airway Chips provided a more favorable environment for *Pseudomonas aeruginosa* growth, which resulted in enhanced secretion of inflammatory cytokines and recruitment of PMNs to the airway.

**Conclusions:** The human CF Airway Chip may provide a valuable preclinical tool for pathophysiology studies as well as for drug testing and personalized medicine.

## 1. Background

Cystic fibrosis (CF) is a genetic disease caused by mutations in the cystic fibrosis transmembrane conductance regulator (CFTR) gene, which encodes a membrane protein with chloride channel function [1]. In the bronchial epithelium, CFTR dysfunction causes contraction of the periciliary volume; accumulation of dense mucus, promoted by hypersecretion of the mucins MUC5AC and MUC5B and by defective mucociliary transport; chronic infection and neutrophilic inflammation, leading to respiratory insufficiency and death [2]. Recurrent infections and non-resolving inflammation are also hallmarks of CF lung disease. The defective innate immune response in CF airways involves toll-like receptor (TLR) signaling [3], reduced height and altered pH of the airway surface liquid layer that impairs the activity of antimicrobial peptides [4,5], segregation of bacteria by a thick MUC5AC-rich mucus, [6] and over-production of proinflammatory cytokines, such as IL-8 [7]. The CF airway milieu also promotes recruitment and pathological conditioning of PMNs [8], which represent key effectors of CF airway infection and inflammation. In addition, CFTR is expressed by vascular endothelial cells [9,10] and its activity is reduced in people with CF, leading to endothelial dysfunction in vitro and in vivo [11–13].

Thus, appropriate tools are needed to study the complex interactions among the epithelial, endothelial and blood components in CF lung airways. This need is particularly urgent in the era of CF therapy with CFTR modulators. Although these drugs have significantly improved the clinical course in CF patients, their long-term impact on lung inflammation still remains unclear. Drug screening platforms with patient-derived cells in which host immune responses to bacterial infection can be studied would therefore also be useful to further develop this pharmacology.

Here, we describe how human organ-on-a-chip (Organ Chip) microfluidic culture technology can be leveraged to create a human Lung Airway Chip lined with primary bronchial epithelial cells (HBE) obtained from CF patients and interfaced with primary human lung microvascular endothelium (CF Airway Chip). We show that this CF Airway Chip recapitulates key features of human CF airways and thus, may provide a new testbed and drug screening tool for researchers and pharmaceutical scientists interested in this life-threating disease.

## 2. Methods

### 2.1. Organ Chip culture

The Organ Chips were cultured in 2-channel microfluidic Chip-S1 devices placed within Pod^®^ holders for culture medium supply and collection using the ZOE^®^ module, as indicated by the manufacturer (Emulate, Inc. Boston). The polydimethylsiloxane (PDMS) chips contain two parallel linear channels separated by a 7 µm pore membrane. Chips were degassed for 30 minutes, activated using the ER1/ER2 reagents following manufacturer’s instructions, washed with HEPES buffer pH 7.4, and coated overnight at 37° C with extracellular matrix (ECM) composed of 50 µg/ml human fibronectin, 50 µg/ml human laminin, and 100 µg/ml collagen I (all from Sigma-Aldrich).

The next day, the chips were washed twice with PBS and primary human lung microvascular endothelial cells (PMVEC) (PromoCell) were introduced in ∼ 20 µL (5 ×10^6^ cells/mL) of EGM-MV2 growth medium (PromoCell) through the inlets of the bottom channel using a micropipette and let them adhere by inverting the chips for 1 hour at 37°C. Chips were then inverted back and healthy or CF human bronchial epithelial cells (HBE) (provided by the University of North Carolina via the CF Foundation; additional healthy HBE cells were purchased from Lonza) were introduced in ∼ 35 µL (3 ×10^6^ cells/mL) of epithelial growth medium (PromoCell) into the top channel. After 2 hours, the channels were rinsed with 200 µL fresh medium and tips were left inserted to prevent drying and allow cell adhesion overnight.

The chips were then placed into the Pods and inserted in the Zoe instrument where they were perfused at a flow rate of 45 µL/hr in both apical and bottom channels with the degassed endothelial or epithelial medium. The chips were maintained under a liquid-liquid Interphase for 4 days before differentiation into a pseudostratified epithelium by shifting to an air-liquid interface (ALI). This was accomplished by perfusing only the lower vascular channel with PneumaCult™-ALI Medium (Stemcell) supplemented with 10 ng/mL VEGF and 1 µg/mL ascorbic acid at 30 µL/hr; the pseudostratified epithelium was established by two weeks in ALI.

### 2.2. Immunostaining

The chips were fixed with 4% paraformaldehyde (PFA) in PBS and stored at 4°C overnight with 200µL tips in the ports. Chips or chip cryosections were permeabilized using 0.2% TRITON-X100 in PBS and blocked with PBS-diluted 5% goat serum for 1 hour at room temperature (RT). Samples were stained overnight at 4°C with primary antibodies: mouse anti-β-tubulin IV (Sigma-Aldrich), rat anti-CC10 (R&D) and rabbit anti-Ck5 (Abcam). The next day, after three washes with PBS, the samples were stained with secondary antibodies: goat anti-mouse/rabbit or rat Alexa-488 or Alexa-546 (Invitrogen), for 1 hour at RT. Samples were washed and stained with 1 µg/ml DAPI for 5 minutes (Abcam) and washed with PBS. For migration studies, chips were stained with PE-conjugated anti CD-31 or Alexa 555-conjugated Anti ZO-1 (both from Invitrogen) to visualize endothelial and epithelial cells, respectively. Images were taken using a Leica SP5 laser scanning confocal immunofluorescence microscope.

### 2.3. Expression of cilia and ciliary beat frequency (CBF)

Fifteen regions of interest along the entire length of the chip epithelial channel were monitored using video imaging on an inverted bright-field microscope. Images were analyzed with a MATLAB script to calculate the CBF and percentage of cell surface area occupied by cilia.

### 2.4. Mucus imaging

Chips were fixed, permeabilized and stained with wheat germ agglutinin-rhodamine and jacalin (both from Vector Laboratories) for 2 hours at 37°C. A razor blade was used to remove 2 mm wide slivers of PDMS from each side of the chip along the length of the channels. The chips were placed onto a glass slide so that the cut side of the chip was facing up and both channels and the intervening membrane could be visualized from the side. Images were taken using an inverted microscope (Zeiss Axio Observer Z1) equipped for dark-field imaging. The areas covered with mucus and fluorescence intensities were analyzed using the Fiji software.

### 2.5. Infection of the Airway Chips

After 11-13 days under ALI, chips were unplugged from the Pods and GFP-labeled *P. aeruginosa* bacteria (20-30 colony forming units; CFU) were inoculated into the apical channel in 35 μL of PBS with calcium and magnesium chloride and incubated for 1 hour. Unattached bacteria were washed out by perfusing additional 100 μL PBS through the apical channel before restoring the ALI. Chips were plugged back into the pods and the flow of ALI medium through the basal vascular channel was restored for additional 24 hours. One day later, the basal outflow was collected for cytokine analysis and bacteria were quantified in live cultures using fluorescence microscopic imaging; the chips were then fixed for additional immunofluorescence staining and imaging. To explore effects of bacterial infection on PMN recruitment and transmigration on-chip, we introduced 20-30 CFU of *P. aeruginosa* on the epithelial side of healthy and CF chips for 8 h, and then PMNs were perfused through the basal channel.

### 2.6. Cytokine quantification

The detection kit “20-Plex Human ProcartaPlex^™^ Panel” (ThermoFisher Scientific) was used to analyze 20 cytokines, including GM-CSF, IFN-α, IFN-γ , IL-1α, IL-1β, IL-4, IL-6, IL-8, IL-10, IL-12p70, IL-13, IL-17A, TNF-α, IP-10, MCP-1, MIP-1 alpha, MIP-1 beta, ICAM-1, CD62E and CD62P. All incubations and washes were executed following the manufacturer’s instructions. The plates were run on FlexMAP3D™ system (Bio-Rad) according to standard protocols.

### 2.7. Immune cell recruitment and migration under flow

PMNs were isolated from fresh blood (< 6 hours collection, RBC, Watertown, MA) using the Ficoll-Paque™ PLUS (Activia) and following manufacturer’s instructions. PMNs were washed with PBS, live stained for 30 minutes at 37° with 10 µM cell tracker green solution (ThermoFisher Scientific) and resuspended at 5 × 10^6^ cells/ mL before being pumped into the chip inlet at 500 µL/hr for 2 hours. Chips were fixed with 4% PFA and stained for immunocytochemistry. The number of PMNs adherent to the endothelial and epithelial layers was measured by analyzing 15 regions of interest per chip (6 chips per condition) using confocal microscopy.

### 2.8. Statistical Analysis

Statistical analyses were performed using GraphPad Prism (GraphPad, San Diego, CA). Results are shown as mean ± standard deviation (SD) or standard error of the mean (SEM), as indicated. Non-parametric data distributions were assessed with the T-test with Mann Whitney, one-way or two-way ANOVA, depending on the experiments; parametric data distribution was assessed with two-tail T-test. Occasional chips that were compromised due to fluid entering the air channel were excluded from the analyses.

## 3. Results

### 3.1. The human CF Airway Chip

We have previously described a method for creating Airway Chips lined by primary human bronchial [14] or bronchiolar [15] epithelial and PMVEC from healthy donors separated by an ECM-coated porous membrane (**Fig. 1A**). In the present study, we created healthy and CF Airway Chips by culturing HBE from healthy subjects or people with CF and compared their phenotypes *in vitro*. Both the healthy and CF epithelium, as well as the endothelium, formed tight monolayers covering the full length of the channels after 12-14 days of culture under ALI (**Fig. 1B,C**). The healthy and CF Airway Chips were lined by a highly differentiated epithelium containing ciliated, basal, club and goblet cells, as detected by ß-tubulin, CK5, CC10, and Muc5AC staining, and ciliated epithelial cells could be seen along the entire length of the channel, when analyzed by immunofluorescence microscopy (**Fig. 1D,E**). The CF Airway Chips contained a higher number of ciliated cells compared to healthy chips after 2 weeks in ALI (**Fig. 1E** and **Supplementary videos 1 and 2**). Quantification of the apical cell surface area covered by cilia confirmed that the epithelium in CF Airway Chips had a significantly larger percentage of its surface covered by ciliated cells than normal epithelium (**Fig. 2A**). In addition, the CF epithelium exhibited a higher CBF compared to the healthy epithelium (**Fig. 2B**), which is consistent with in vivo data [16].

**Figure 1.**
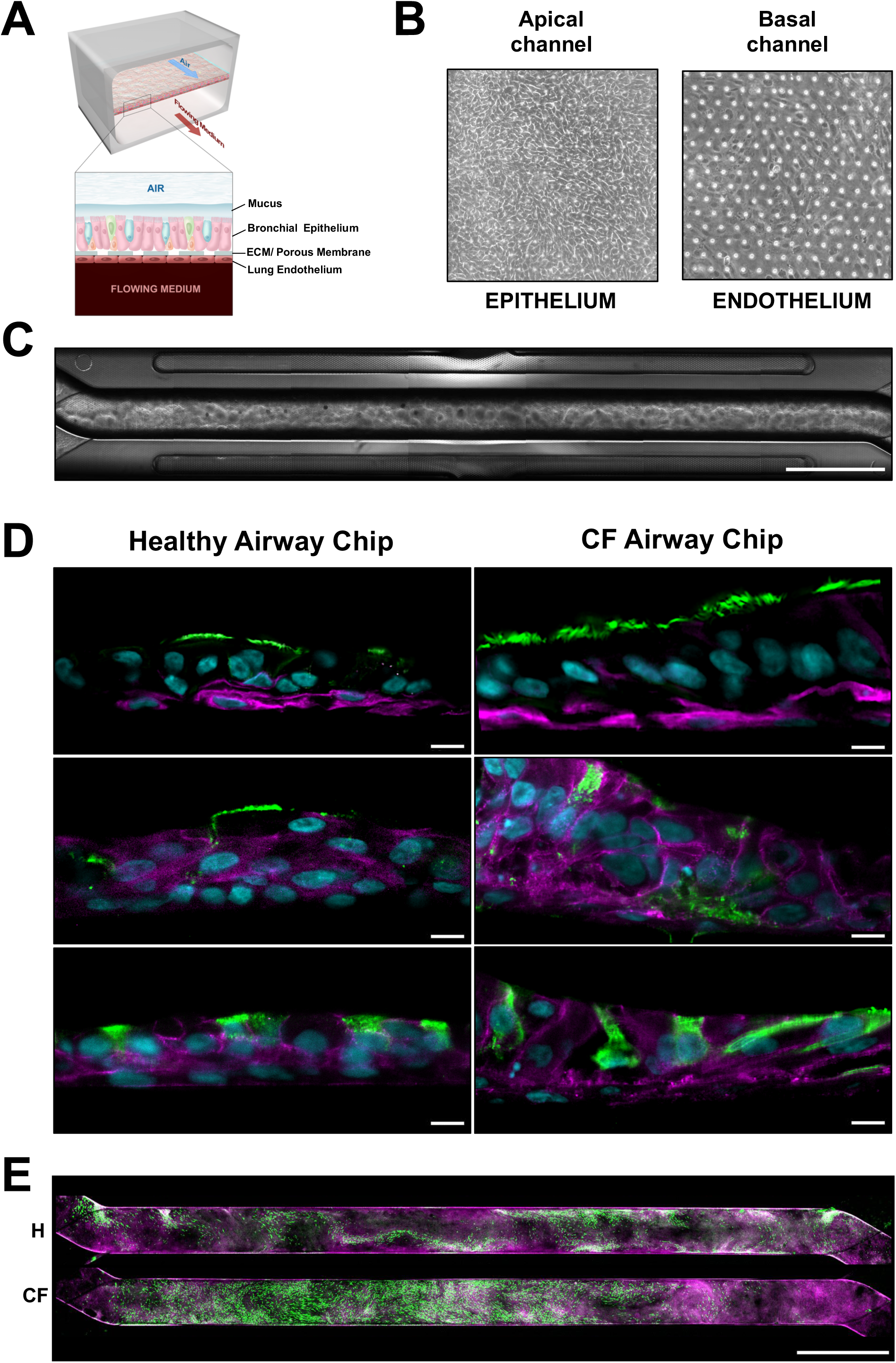
Human healthy and CF Airway Chips. (**A**) Diagram of the Airway Chip showing the pseudostratified bronchial epithelium cultured under an ALI on the top surface of the porous membrane separating the upper epithelial channel and the lower vascular channel that contains human lung PMVECs adherent to the bottom of the same membrane and exposed to dynamic fluid flow. (**B**) Representative phase contrast images of bronchial epithelial cells forming a monolayer on the top surface of the porous membrane (left) and the endothelium on the lower surface of the same membrane (right). The regular array of small white dots visible at right are the 7 μm pores in the membrane. (**C**) Low magnification phase contrast microscopic view of the entire length of the chip viewed from above after 12 days of growth of the epithelium under an ALI (channel width is 1 mm; bar, 2 mm). (**D**) Representative immunofluorescence confocal microscopic images of vertical cross sections of healthy and CF Aiway Chips, showing ciliated cells expressing β-Tubulin IV (green) and basal cells expressing CK5 (magenta) in the top view, goblet cells positive for MUC5AC (green) at the bottom and Clara cells expressing CC-10 (green) in the center view (bar, 10 μm). (**E**) Representative low magnification, immunofluorescence confocal micrographs of the entire length of the epithelial channel in healthy (H) versus CF Airway Chips (CF) stained for F-actin (phalloidin; magenta) and β-tubulin IV (green) to visualize ciliated cells (bar, 2 mm).

**Figure 2.**
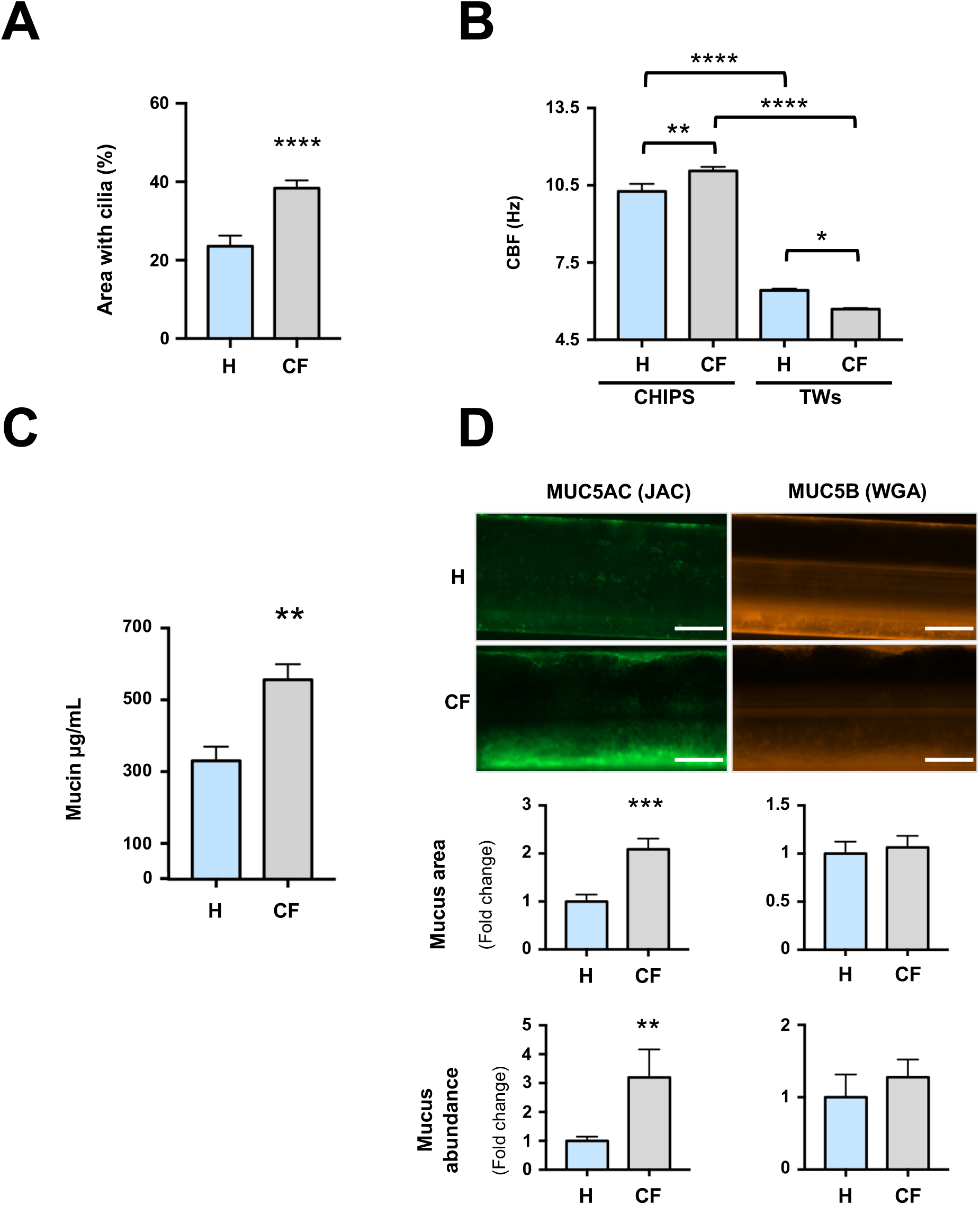
Cilia and mucus in healthy and CF Airway Chips. (**A**) The apical surface area of the epithelium expressing cilia in healthy (H) versus CF Airway (CF) Chips. Data are expressed as % total surface area per field. (**B**) Ciliary beat frequency (CBF) in Hz measured in healthy versus CF Airway Chips or Transwells (TWs). (**C**) Total mucin harvested from the epithelium after 7 days of culture under ALI from healthy versus CF Airway Chips, as measured by Alcian Blue staining. (**D**) Representative side view images of healthy and CF Airway Chips stained for MUC5AC with jacalin (JAC) or for MUC5B with wheat germ agglutinin (WGA) (right), and respective histograms below showing quantification of the area covered by the signal, and densitometric analysis (bar, 500 μm). All data represent mean ± SD obtained from at least 2 separate experiments; n = 22-30 ROIs from 2 different chips or TWs for **A** and **B**; n = 6 biological replicates (3 chips from 2 healthy donors and 2 CF donors each) in **C**; n = 4 chip replicates for **D**; ^****^ p<0.0001, ^***^ p<0.001, ^**^ p<0.01, ^*^ p <0.05.

Importantly, this mimicry of high in vivo-like CBFs could not be detected when epithelial cells were cultured in TWs (**Fig. 2B**).

Low-magnification phase contrast microscopic analysis also suggested that mucus accumulated above the epithelium (**Fig. 1C**). We therefore collected apical washes from the chip epithelium-lined channels at day 7 of ALI and quantified mucin abundance by Alcian blue staining. These studies confirmed that the CF Airways Chips displayed more abundant mucin accumulation compared to healthy chips (**Fig. 2C**). The CF chips also exhibited a greater percentage of area covered by MUC5AC, and mucus abundance, whereas there was no significant difference in the amount of MUC5B (**Fig. 2D**) when stained with fluorescent jacalin and wheat germ agglutinin (WGA) lectins, respectively. Thus, the CF Airway Chips recapitulated the increased ciliation, higher CBF, and enhanced mucus accumulation observed in the airways of CF patients [16,17].

### 3.2. CF Airway Chips exhibit baseline inflammation and enhanced neutrophil recruitment

CF lungs exhibit enhanced inflammation [18] and we similarly observed a significant increase in the pro-inflammatory cytokine, IL-8, with decreased levels of IP-10, GM-CSF and MIP1-, and no differences in IL-6, in the vascular effluents of CF Chips compared to the healthy ones after 2 weeks in ALI (**Fig. 3A**). We then explored immune cell behavior in the pro-inflammatory microenvironment of the CF Airway Chip by flowing PMNs through its vascular channel and comparing the response to that produced in healthy Airway Chips. PMNs spontaneously adhered to PMVECs (**Fig. 3B,C**) and migrated through the ECM-coated pores of the central membrane to reach the epithelial surface in both healthy and CF Chips (**Fig. 3D,E**).

**Figure 3.**
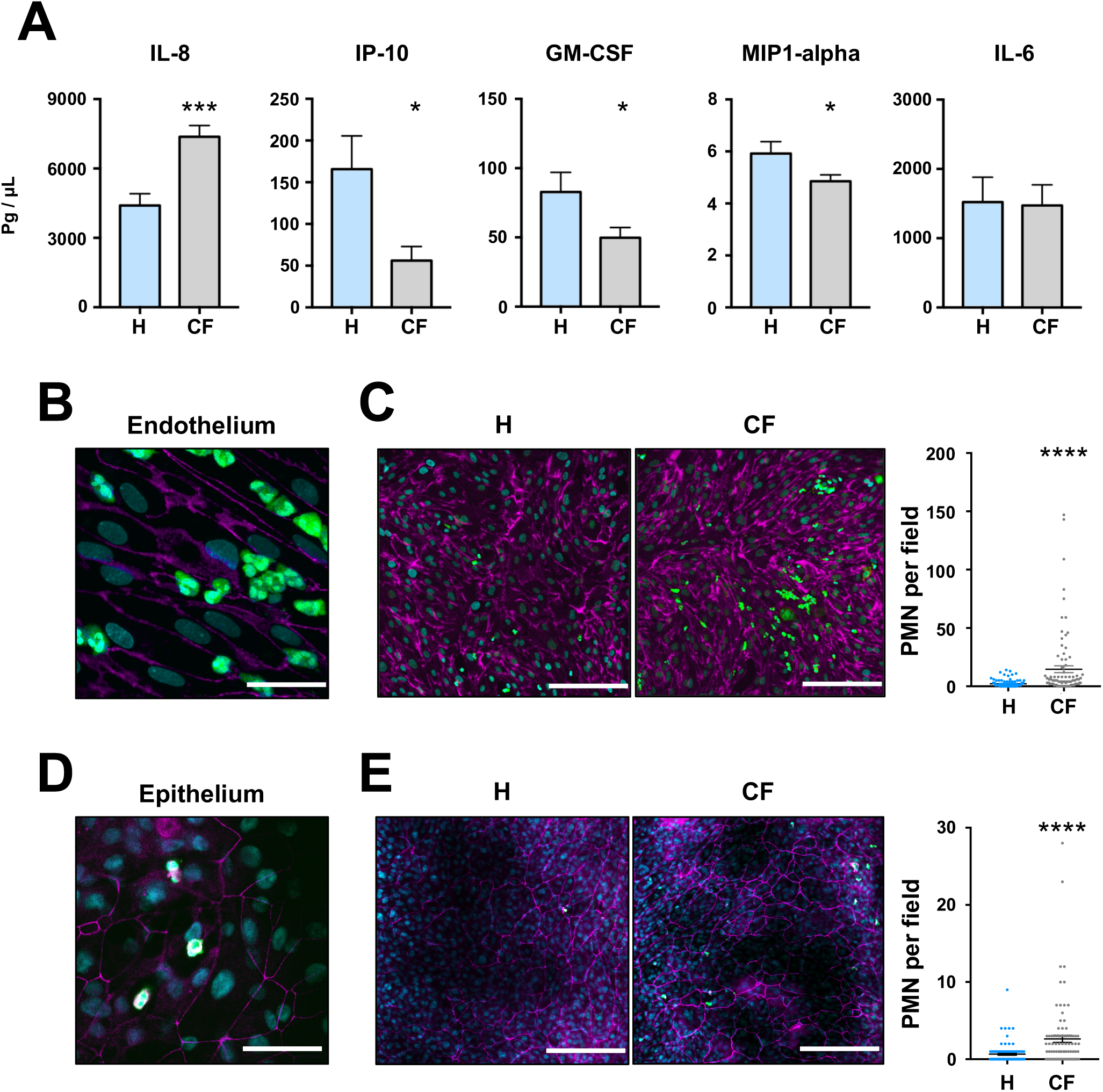
Baseline inflammatory phenotype in healthy versus CF chips under sterile conditions. (**A**) Profile of inflammatory cytokines, IL-8, IP-10, GM-CSF, MIP1-alpha, and IL-6, quantified in the basal vascular outflow of healthy and CF Airway Chips after 10 days in culture. High (**B**,**D**) and low (**C**,**E**) magnification confocal fluorescence microscopic images showing PMNs (green) adherent to the CD31 expressing endothelium (magenta) (**B**,**C**) and transmigrated to the ZO-1 containing epithelium in the upper channel (**D**,**E**) in healthy (H) versus CF Airway (CF) Chips 2 hours after being flowed through the vascular channel (bar, 200 μm). The graphs at the right in **C** and **E** show quantification of the PMN recruited per high power field to the surface of the endothelium and epithelium, respectively. All data represent mean ± SEM; n = 6 biological chip replicates from two different experiments in **A-E;** 15 fields per chip were used for quantification in **C** and **E**; ^****^ p<0.0001, ^***^ p<0.001, ^**^ p<0.01, ^**^ p <0.05.

However, consistent with the enhanced IL-8 release, we observed significantly increased PMN adhesion and endothelial to epithelial transmigration in the CF chips (**Fig. 3C,E**), which is again reminiscent of the increased immune cell infiltration observed in CF lungs.

### 3.3. Responses of the CF Airway Chip to *P. aeruginosa* infection

*P. aeruginosa* frequently colonizes CF airways [19], and so to mimic infection, we inoculated the chips with GFP-labeled *P. aeruginosa* bacteria. Twenty-four hours post infection, the epithelium appeared intact in both healthy and CF Chips (**Fig. 4A**), whereas large clusters of GFP-labeled bacteria could be detected in the mucus layer closely apposed to the underlying epithelium (**Fig 4B**). Fluorescence densitometric quantitation showed a higher number of bacteria in the CF Airway Chips compared to the healthy chips (**Fig. 4C**), which is consistent with the reported enhanced growth of *P. aeruginosa* in the CF mucus *in vivo* [20]. Indeed, when the mucus extracted from the Airway Chips was used as a culture medium for *P. aeruginosa*, we observed more pronounced bacterial growth in the mucus from the CF Chips, whereas no difference was detected when similar studies were carried out using mucus from static TW cultures (**Fig. 4D**). Importantly, bacterial infection also resulted in a significant increase in IL-6, TNF- and GM-CSF levels in the vascular effluents of both healthy and CF Airway Chips, whereas there was no further increase in IL-8 levels (**Fig. 4E**). Interestingly, PMN endothelial adhesion was more pronounced in CF Airway Chips with *P. aeruginosa* infection **(Fig. 5B)**, but we could not detect a significant difference in the number of PMNs transmigrated to the epithelium (**Fig. 5C**).

**Figure 4.**
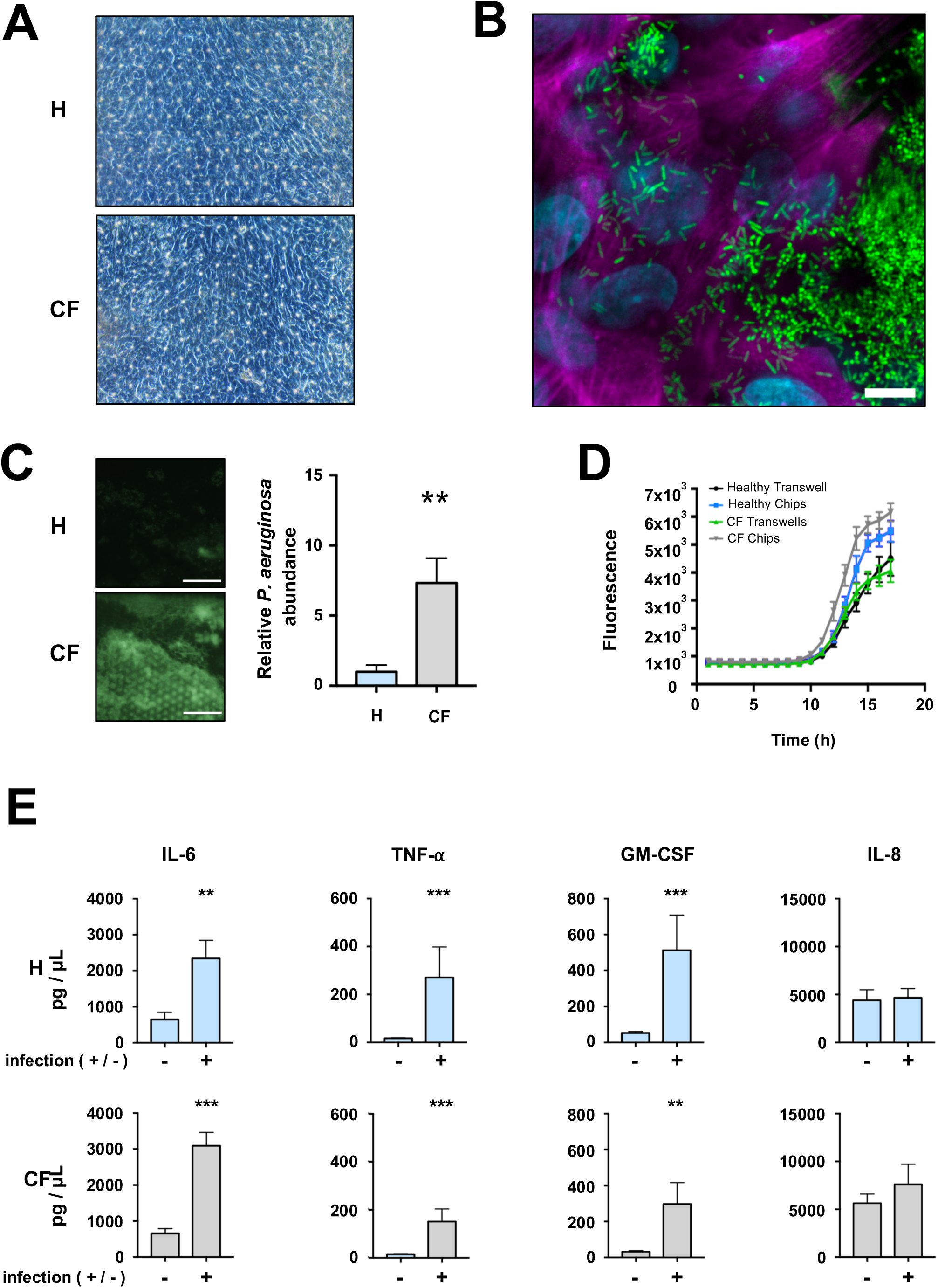
*P. aeruginosa* infection of healthy and CF chips. (**A**) Brightfield microscopic images of epithelium in healthy (H) versus CF Airway (CF) Chips 24 hours after infection with *P. aeruginosa*. (**B**) Fluorescence confocal micrograph showing clusters of GFP-*P. aeruginosa* bacteria growing on the airway epithelium 24 hours after infection (bar, 10 μm). (**C**) Representative images of GFP-*P. aeruginosa* (left) in healthy and CF Airway Chips at lower magnification (bar, 200 m) and quantification by densitometric analysis 24 hour post infection (right). (**D**) Bacterial growth curve measured by quantification of fluorescence intensity when the GFP-bacteria were grown in mucus collected from healthy or CF Airway Chips versus Transwells. (**E**) Quantification of cytokine present in the vascular effluent 24 hours post-infection in infected and non-infected healthy and CF Airway Chips. Data represent mean ± SEM; n = 4 biological chip replicates for **C**, n = 4-6 for **D**, and n ≥ 6 for **E**. Two-way Anova was used for statistical analysis of panel **D** and it revealed statistical significance between CF Airway Chips compared to healthy chips [10 hr (^*^), 11 hr (^**^), and 12 hr (^**^)]; CF Airway Chips versus healthy TWs [12 hr (^**^), 13-17 hr (^****^)]; CF Chips versus CF TWs [12 hr (^*^), 13-17 hr (^****^)]; healthy chips versus healthy TWs [14 hr (^***^), 15 hr (^****^), 16-17 hr (^**^)]; and healthy chips vs CF TWs [14 hr (*), 15 hr (^***^), 16-17 hr (^****^)]. ^****^ p<0.0001, ^***^ p<0.001, ^**^ p<0.01, ^*^ p <0.05.

**Figure 5.**
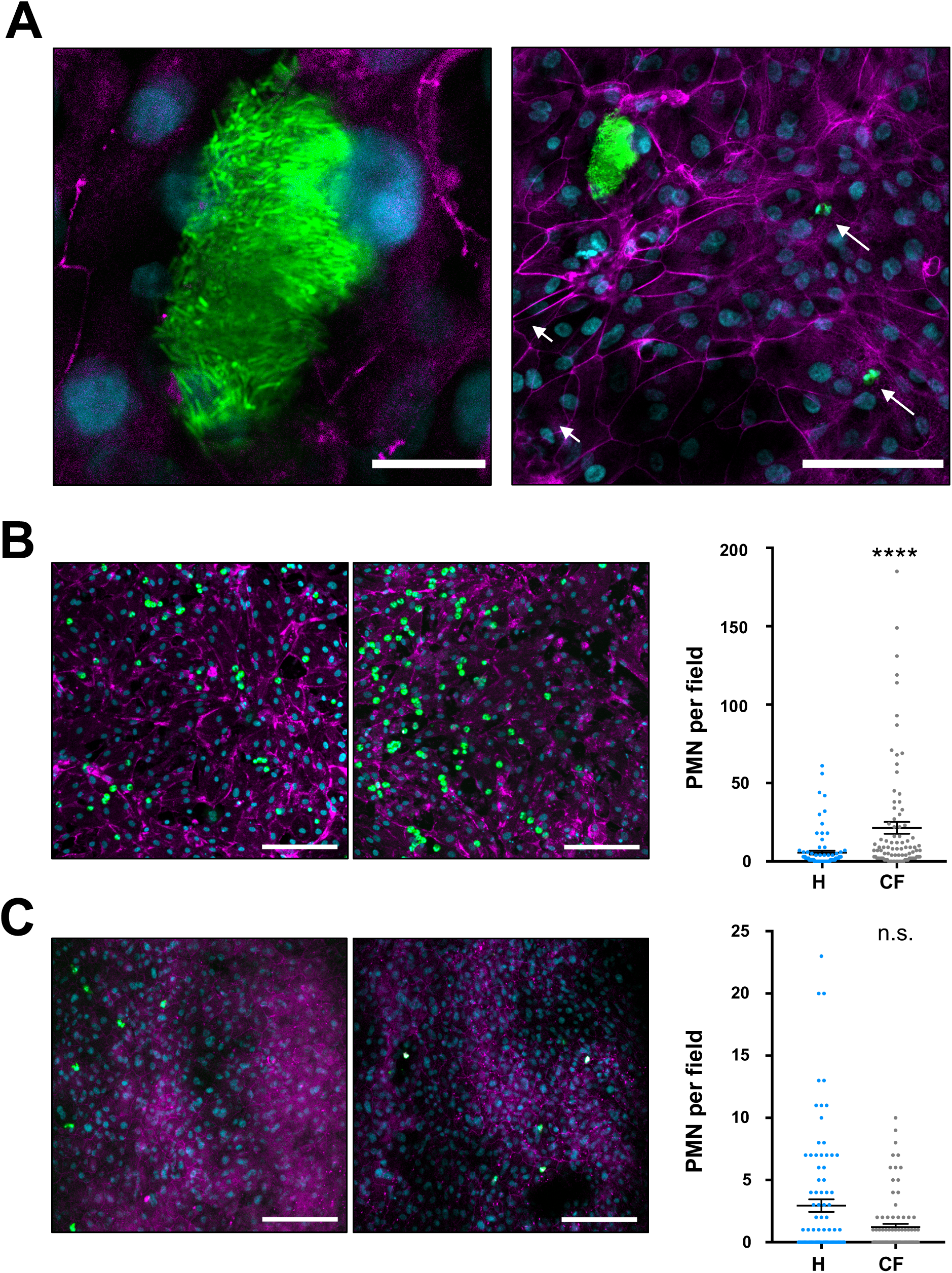
PMN adhesion and transmigration in healthy versus CF Airway Chips infected with *P. aeruginosa*. (**A**) High (left) and lower (right) magnification confocal fluorescence microscopic views of a dense GFP-*P. aeruginosa* colony growing on the epithelium of an infected CF Airway Chip (left) (bars, 20 μm and 100 μm). Arrows at right indicate PMNs (green) that are approaching the bacterial colony after extravasation and transmigration through the phalloidin-stained epithelium (magenta). Lower magnification confocal microscopic images (left) and corresponding quantification (right) of PMNs (green) adherent to the CD31-expressing endothelium (magenta) **(B**) and ZO-1 expressing epithelium (**C**) in healthy (H) versus CF Airway (CF) Chips infected with *P. aeruginosa* 2 hours after being flowed through the vascular channel of the chips (bar, 200 μm). Data represent mean ± SEM of n = 6 biological chip replicates from 2 experiments), 15 fields per chip were used for quantification. ^****^ p<0.0001.

## 4. Discussion

Research on CF lung disease has been limited by the relatively low translatability of data obtained with murine models because the pathological features of this disease in mice do not precisely match those in humans [21]. There is therefore a great need for human relevant preclinical models that more closely recapitulate clinical features of the disease. Here, we show that human Organ Chip microfluidic culture technology can be used to create a CF Airway Chip populated with human primary epithelial bronchial cells obtained from patients with CF or non-CF donors. The epithelium is grown under ALI and interfaced with PMVECs that are exposed to dynamic fluidic flow, which permits co-culture of clinically relevant pathogens, such as *P. aeruginosa*, in the air space, as well as circulating immune cells in the vascular channel. Using this chip, we were able to reproduce key features of human CF airways, including higher ciliary activity and mucus that supported greater bacterial growth, which was not replicated in static TW cultures. We also observed accumulation of a thick viscous mucus layer, reminiscent of human CF airways, and the MUC5AC:MUC5B ratio was similar to that reported for mucus from patients with CF [6,21]. Thus, the CF chip recapitulates many of the key features of the airway dysfunction present in people with CF [22,23].

Non-resolving neutrophilic respiratory inflammation is another hallmark of CF [18] as CF airways commonly display massive PMN infiltration and release high amounts of pro-inflammatory cytokines [7,18]. Importantly, these characteristics were also reproduced in our preclinical model as uninfected CF Airway Chips exhibited enhanced secretion of cytokines, such as the potent PMN chemo-attractant IL-8 [24], and increased adhesion of circulating PMNs as well as their transmigration from the vascular channel, through the tissue-tissue interface, and into the epithelium-lined apical channel of the chip compared to healthy Airway Chips. These results are consistent with the concept that CFTR loss-of-function confers a pro-inflammatory phenotype, independently from infection [11] and that CF airway can condition circulating PMNs [8].

Our results showing that mucus collected from the airway channel of CF Airway Chips proves a more favorable environment for growth of *P. aeruginosa* bacteria than mucus produced either by healthy Airway Chips or CF cells cultured in static TW inserts, demonstrate another novelty ability of this system to replicate the CF airway microenvironment and associated pathophysiology. This finding is consistent with the observation that CF patient sputum supports higher density growth of *P. aeruginosa* when used as a substrate compared to sputum from healthy people [25]. In addition, CF Airway Chips exposed to *P. aeruginosa* released high levels of IL-6, as well as TNF-α and GM-CSF, which is in agreement with the higher IL-6 levels observed in past *in vitro* studies with CF HBE cell cultures infected with *P. aeruginosa*. These findings are also consistent with the negative correlation observed between TNF-α levels in CF patients’ sputum after disease exacerbations and their respiratory indexes, FEV1 and FVC [26].

TNF-α increases endothelial expression of ICAM-1 and stimulates PMN adhesion to the endothelium, in addition to enhancing expression of the IL-6 receptor [27,28]. Indeed, PMN adherence to the endothelium was enhanced in response to *P. aeruginosa* infection, especially in CF Airway Chips, and this scaled with higher TNF-α levels produced under these conditions. The accompanying increase of the GM-CSF chemokine, which stimulates neutrophil recruitment and enhances their killing activity [29], also could contribute to the increased recruitment we observed. However, transmigration to the CF epithelium under *P. aeruginosa* infection was not different from that observed in the healthy epithelium. This latter finding might be related to the observation that the release of the main PMN chemoattractant, IL-8, was not different between healthy and CF chips upon infection with *P. aeruginosa*.

Although the results shown here that were obtained with the CF Airway Chip reproduce many features of the diseased lung airway of CF patients, this model has room to be improved. For example, PMNs and PMVECs obtained from non-CF individuals were used in these chips, and even better recapitulation of clinical phenotype might be able to be obtained using all cell types from the CF patients, or even from the same patient. Additional cell types, such as pulmonary macrophages and lung fibroblasts, are also missing from the model, and they could be added in the future as well. Indeed, the pathogenetic relevance of the airway connective tissue has been recently documented in a 3D CF stromal lung model [30]. It is possible that the absence of some or all of these CF patient-derived cells could contribute to the explanation of why we did not observe a difference in the number of PMNs that transmigrated to the epithelial channel between healthy and CF Airway Chips infected with *P. aeruginosa*.

In conclusion, the human CF Airway Chip reproduced for the first time, and with high fidelity, many of the structural, biochemical, and pathophysiological features of the human CF lung airway and its response to pathogens and circulating immune cells *in vitro*. This microfluidic Organ Chip model that is lined by human patient-derived cells opens the possibility to create personalized preclinical models for mechanistic studies as well as drug testing and discovery that could help to advance the CF research field and accelerate the delivery of more effective therapeutics to patients.

## Data Availability

All data are included in the submission and will be made available upon request

## Acknowledgments

This work was supported by the Cystic Fibrosis Foundation, the Wyss Institute for Biologically Inspired Engineering at Harvard University, and Programma Operativo Nazionale Ricerca e Innovazione (D56C19000330005).

## Author contributions

R. Plebani and R. Potla designed the study with D.E.I. and performed most experiments with culture and analytical assistance from M.S., H.B., S.G., and C.B.; Z.I. performed the mucin analysis and A.J. carried out the Luminex assays; R.N.T. and M.C. cultured the GFP *P. aeruginosa* and assisted in the analysis. P.J., S.G., R.P-B., M.R., and D.E.I. supervised and managed the program, reviewed data, and assisted in the preparation of the manuscript along with all other authors.

## REFERENCES

[1] Riordan JR, Rommens JM, Kerem B, Alon N, Rozmahel R, Grzelczak Z, et al. Identification of the cystic fibrosis gene: cloning and characterization of complementary DNA. Science 1989;245:1066–73. https://doi.org/10.1126/science.2475911.

[2] Stoltz DA, Meyerholz DK, Welsh MJ. Origins of Cystic Fibrosis Lung Disease. N Engl J Med 2015;372:351–62. https://doi.org/10.1056/NEJMra1300109.

[3] Greene CM, Carroll TP, Smith SGJ, Taggart CC, Devaney J, Griffin S, et al. TLR-Induced Inflammation in Cystic Fibrosis and Non-Cystic Fibrosis Airway Epithelial Cells. The Journal of Immunology 2005;174:1638–46. https://doi.org/10.4049/jimmunol.174.3.1638.

[4] Birket SE, Chu KK, Liu L, Houser GH, Diephuis BJ, Wilsterman EJ, et al. A functional anatomic defect of the cystic fibrosis airway. Am J Respir Crit Care Med 2014;190:421–32. https://doi.org/10.1164/rccm.201404-0670OC.

[5] Abou Alaiwa MH, Reznikov LR, Gansemer ND, Sheets KA, Horswill AR, Stoltz DA, et al. pH modulates the activity and synergism of the airway surface liquid antimicrobials β-defensin-3 and LL-37. Proc Natl Acad Sci USA 2014;111:18703–8. https://doi.org/10.1073/pnas.1422091112.

[6] Fernández-Blanco JA, Fakih D, Arike L, RodrÍguez-Piñeiro AM, MartÍnez-Abad B, Skansebo E, et al. Attached stratified mucus separates bacteria from the epithelial cells in COPD lungs. JCI Insight 2018;3. https://doi.org/10.1172/jci.insight.120994.

[7] Tabary O, Zahm JM, Hinnrasky J, Couetil JP, Cornillet P, Guenounou M, et al. Selective Up-Regulation of Chemokine IL-8 Expression in Cystic Fibrosis Bronchial Gland Cells in Vivo and in Vitro. The American Journal of Pathology 1998;153:921–30. https://doi.org/10.1016/S0002-9440(10)65633-7.

[8] Forrest OA, Ingersoll SA, Preininger MK, Laval J, Limoli DH, Brown MR, et al. Frontline Science: Pathological conditioning of human neutrophils recruited to the airway milieu in cystic fibrosis. J Leukoc Biol 2018;104:665–75. https://doi.org/10.1002/JLB.5HI1117-454RR.

[9] Tousson A, Van Tine BA, Naren AP, Shaw GM, Schwiebert LM. Characterization of CFTR expression and chloride channel activity in human endothelia. American Journal of Physiology-Cell Physiology 1998;275:C1555–64. https://doi.org/10.1152/ajpcell.1998.275.6.C1555.

[10] Plebani R, Tripaldi R, Lanuti P, Recchiuti A, Patruno S, Di Silvestre S, et al. Establishment and long-term culture of human cystic fibrosis endothelial cells. Lab Invest 2017;97:1375–84. https://doi.org/10.1038/labinvest.2017.74.

[11] Totani L, Plebani R, Piccoli A, Di Silvestre S, Lanuti P, Recchiuti A, et al. Mechanisms of endothelial cell dysfunction in cystic fibrosis. Biochim Biophys Acta Mol Basis Dis 2017;1863:3243–53. https://doi.org/10.1016/j.bbadis.2017.08.011.

[12] Romano M, Collura M, Lapichino L, Pardo F, Falco A, Chiesa PL, et al. Endothelial perturbation in cystic fibrosis. Thromb Haemost 2001;86:1363–7.

[13] Poore S, Berry B, Eidson D, McKie KT, Harris RA. Evidence of vascular endothelial dysfunction in young patients with cystic fibrosis. Chest 2013;143:939–45. https://doi.org/10.1378/chest.12-1934.

[14] Si L, Bai H, Rodas M, Cao W, Oh CY, Jiang A, et al. A human-airway-on-a-chip for the rapid identification of candidate antiviral therapeutics and prophylactics. Nat Biomed Eng 2021. https://doi.org/10.1038/s41551-021-00718-9.

[15] Benam KH, Villenave R, Lucchesi C, Varone A, Hubeau C, Lee H-H, et al. Small airway-on-a-chip enables analysis of human lung inflammation and drug responses in vitro. Nat Methods 2016;13:151–7. https://doi.org/10.1038/nmeth.3697.

[16] Alikadic S, Horak F, Frischer T, Nachbaur E, Renner S, Gruber S. Ciliary beat frequency in nasal and bronchial epithelial cells in patients with cystic fibrosis. European Respiratory Journal 2011;38.

[17] Kirkham S, Sheehan JK, Knight D, Richardson PS, Thornton DJ. Heterogeneity of airways mucus: variations in the amounts and glycoforms of the major oligomeric mucins MUC5AC and MUC5B. Biochem J 2002;361:537–46.

[18] Kruger P, Saffarzadeh M, Weber ANR, Rieber N, Radsak M, von Bernuth H, et al. Neutrophils: Between host defence, immune modulation, and tissue injury. PLoS Pathog 2015;11:e1004651. https://doi.org/10.1371/journal.ppat.1004651.

[19] Emerson J, Rosenfeld M, McNamara S, Ramsey B, Gibson RL. Pseudomonas aeruginosa and other predictors of mortality and morbidity in young children with cystic fibrosis. Pediatr Pulmonol 2002;34:91–100. https://doi.org/10.1002/ppul.10127.

[20] Isopi E, Mattoscio D, Codagnone M, Mari VC, Lamolinara A, Patruno S, et al. Resolvin D1 Reduces Lung Infection and Inflammation Activating Resolution in Cystic Fibrosis. Front Immunol 2020;11. https://doi.org/10.3389/fimmu.2020.00581.

[21] Henderson AG, Ehre C, Button B, Abdullah LH, Cai L-H, Leigh MW, et al. Cystic fibrosis airway secretions exhibit mucin hyperconcentration and increased osmotic pressure. J Clin Invest 2014;124:3047–60. https://doi.org/10.1172/JCI73469.

[22] Henke MO, John G, Germann M, Lindemann H, Rubin BK. MUC5AC and MUC5B mucins increase in cystic fibrosis airway secretions during pulmonary exacerbation. Am J Respir Crit Care Med 2007;175:816–21. https://doi.org/10.1164/rccm.200607-1011OC.

[23] Xie Y, Ostedgaard L, Abou Alaiwa MH, Lu L, Fischer AJ, Stoltz DA. Mucociliary Transport in Healthy and Cystic Fibrosis Pig Airways. Ann Am Thorac Soc 2018;15:S171–6. https://doi.org/10.1513/AnnalsATS.201805-308AW.

[24] Richman-Eisenstat JB, Jorens PG, Hebert CA, Ueki I, Nadel JA. Interleukin-8: an important chemoattractant in sputum of patients with chronic inflammatory airway diseases. American Journal of Physiology-Lung Cellular and Molecular Physiology 1993;264:L413–8. https://doi.org/10.1152/ajplung.1993.264.4.L413.

[25] Palmer KL, Mashburn LM, Singh PK, Whiteley M. Cystic fibrosis sputum supports growth and cues key aspects of Pseudomonas aeruginosa physiology. J Bacteriol 2005;187:5267–77. https://doi.org/10.1128/JB.187.15.5267-5277.2005.

[26] Colombo C, Costantini D, Rocchi A, Cariani L, Garlaschi ML, Tirelli S, et al. Cytokine levels in sputum of cystic fibrosis patients before and after antibiotic therapy. Pediatr Pulmonol 2005;40:15–21. https://doi.org/10.1002/ppul.20237.

[27] Gamble JR, Harlan JM, Klebanoff SJ, Vadas MA. Stimulation of the adherence of neutrophils to umbilical vein endothelium by human recombinant tumor necrosis factor. Proc Natl Acad Sci U S A 1985;82:8667–71.

[28] Romano M, Sironi M, Toniatti C, Polentarutti N, Fruscella P, Ghezzi P, et al. Role of IL-6 and its soluble receptor in induction of chemokines and leukocyte recruitment. Immunity 1997;6:315–25. https://doi.org/10.1016/s1074-7613(00)80334-9.

[29] Spiekermann K, Roesler J, Emmendoerffer A, Elsner J, Welte K. Functional features of neutrophils induced by G-CSF and GM-CSF treatment: differential effects and clinical implications. Leukemia 1997;11:466–78. https://doi.org/10.1038/sj.leu.2400607.

[30] Mazio C, Scognamiglio LS, De Cegli R, Galietta LJV, Di Bernardo D, Casale C, et al. Intrinsic Abnormalities of Cystic Fibrosis Airway Connective Tissue Revealed by an In Vitro 3D Stromal Model. Cells 2020;9:1371. https://doi.org/10.3390/cells9061371.

