## Supplementary_Figure_1 for "Modeling Pulmonary Cystic Fibrosis in a Human Lung Airway-on-a-chip"

### Supplementary Figures

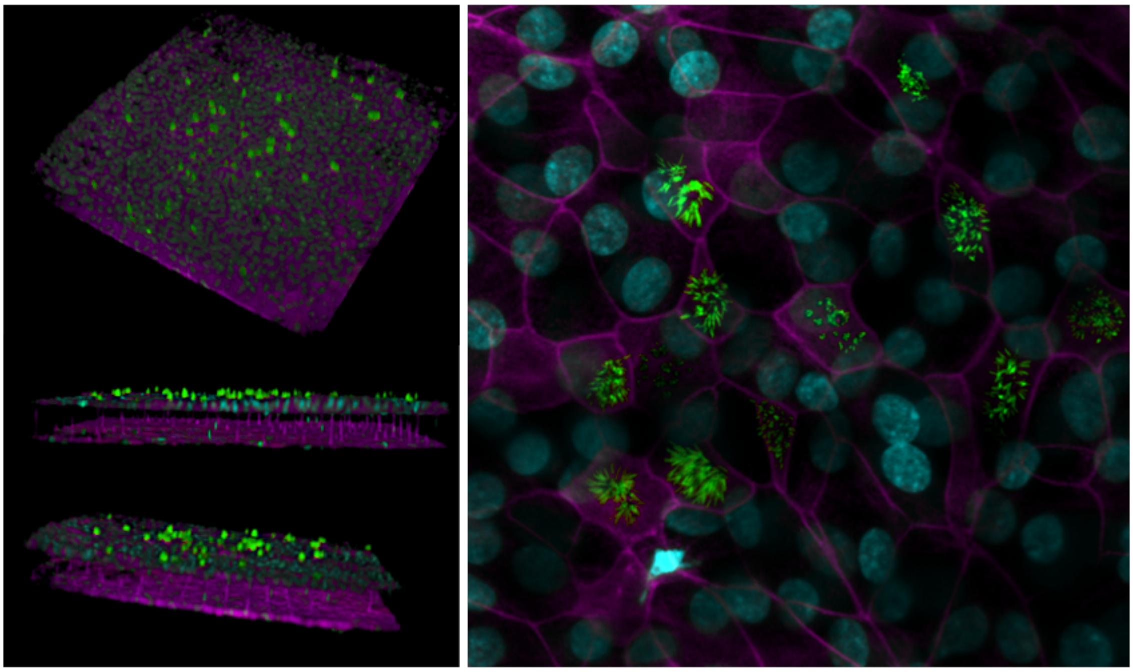

**Supplementary Figure S1.** Confocal immunofluorescence micrographic 3D reconstruction of the interface between the pseudostratified epithelium and endothelium in the human Airway Chip when viewed from above and the side (left) and a higher magnification view from above of the epithelium stained for β-tubulin IV (green) and ZO-1 (magenta) to show the ciliated epithelial cells cultured on top of the porous PDMS membrane and for CD31 to visualize the endothelium below (bar, 50 μm).
